# Berry Supplement-Modified Human Microbiome Improves Immunotherapy Response in Mouse Models

**DOI:** 10.1101/2025.01.16.25320666

**Authors:** Shiva Jahanbahkshi, Amna Bibi, Rebecca Hoyd, Caroline Dravillas, Nyelia Williams, Shiqi Zhang, Aaditya Pallerla, Shankar Suman, Joseph Amann, Mounika Goruganthu, Tamio Okimoto, Yangyang Liu, Marisa A. Bittoni, Ni Shi, Alvin Anand, Bailey Conrad, Lane Nevers, Kristen Heitman, Maxine Webb, Elizabeth M Grainger, Madison Grogan, Christian Quiles, Tong Chen, Carolyn J. Presley, Lang Li, Patrick Bradley, Yael Vodovotz, David P. Carbone, Steven K. Clinton, Jiangjiang Zhu, Daniel Spakowicz

**Affiliations:** Department of Food Science and Technology, College of Food, Agriculture, and Environmental Sciences, The Ohio State University; Columbus, OH, USA; Division of Medical Oncology, Department of Internal Medicine, The Ohio State University Comprehensive Cancer Center; Columbus, OH, USA; Department of Human Sciences and Comprehensive Cancer Center, The Ohio State University College of Medicine; Columbus, OH, USA; Pelotonia Institute for Immuno-Oncology, The Ohio State University Comprehensive Cancer Center; Columbus, OH, USA; The Ohio State University College of Medicine, Columbus, OH, USA; Department of Microbiology, The Ohio State University; Columbus, OH, USA; Department of Biomedical Informatics, The Ohio State University College of Medicine; Columbus, OH, USA

## Abstract

Cancer outcomes have improved with immune checkpoint inhibitor (ICI) treatment; however, less than half of tumors respond. Emerging data show that some responses to ICI depend on the host’s microbiome. Here, we explore a dietary intervention to modify the microbiome and determine the response to ICIs. Following pre-clinical studies showing the benefit of black raspberries, we conducted a human intervention trial called the BEWELL Study (NCT04267874). The intervention increased the abundance of pro-ICI-response microbes (PIRMs). In mouse models, participants’ post-dietary intervention stool led to smaller tumor volumes following treatment with ICIs than pre-dietary intervention samples. One PIRM, *Blautia obeum*, was sufficient to improve ICI response. These results suggest that black raspberry nectar can modify the human gut microbiome to promote an improved response to ICIs.

## Introduction

Cancer is a major public health problem worldwide and the second leading cause of death in the United States (*1*). Immune checkpoint inhibitors (ICIs) have significantly improved clinical outcomes in various cancers (*2*), particularly monoclonal antibodies targeting PD-1, PD-L1, or CTLA-4, which block T cell activity. However, despite their success, responses to ICI treatment remain heterogeneous, with 0–80% (median 30%) of tumors decreasing in size following treatment (*3*, *4*). There is an urgent need to increase the fraction of patients whose tumors respond to ICI treatment.

One area of emerging interest in improving ICI efficacy is the gut microbiome. Pre-treatment stool samples of cancer patients can predict tumor response to ICIs (*5–7*), and fecal transplants can resensitize treatment-resistant tumors (*8*). Many efforts are underway to understand how microbes affect ICI response and to modify the microbiome to improve ICI treatment outcomes. Dietary interventions are particularly attractive, as they are accessible and patient-controlled, but have limited evidence regarding their ability to alter the microbiome on treatment-relevant timescales or with sufficient specificity (*9*).

Here, we investigated a high-polyphenol, whole-food intervention (black raspberries (BRB)) for its ability to modify the gut microbiome to improve ICI treatment response. We demonstrated that BRB powder increases the abundance of pro-ICI-response microbes (PIRMs) in mouse models. In a human clinical trial, we observed that four weeks of 354 mL BRB nectar per day altered the gut microbiomes of study participants, largely by increasing the abundance of microbes in the family *Lachnospiraceae*. We further discovered that the BRB-modified microbiomes improve treatment outcomes in an ICI-response mouse model, relative to those study participants’ pre-intervention samples. Finally, we demonstrated isolated *Lachnospiraceae* strains are sufficient to improve ICI response in mice when supplemented into a human’s pre-BRB gut microbiome. Explorations of the mechanism suggest a role for altered L-carnitine signaling.

## Results

### Preclinical model exploration for food products that enrich gut Pro-Immunotherapy Response Microbes (PIRMs)

We began testing food supplements in mouse models to find dietary interventions to enrich microbes with a literature precedence for affecting ICI response in humans, labeled here as PIRMs. We fed rats a standard synthetic diet (AIN-76A) alone or supplemented with 5% whole BRB powder *ad libitum* for 35 weeks (**Fig. 1A**). We collected stool at the end of the study to test for differences in the gut microbiomes by diet. Clustering by total 16S abundance revealed no separation between the two dietary groups (p-value = 0.096). The black raspberry group showed tighter clustering than the control group, suggesting uniform alterations with the diet (p-value = 0.344) (**Fig. 1B**). We tested for specific taxonomic changes, aggregating to every taxonomic level and correcting for multiple hypothesis tests. We observed significant enrichment of the PIRM *Akkermansia* in the black raspberry group (log2fc = 5.29, adjusted p-value = 0.049), along with its family (formerly *Verrucomicrobiaceae,* now *Akkermansiaceae*) and order (*Verrucomicrobiales*). The cross-taxonomy enrichment demonstrates the selective enrichment of *Akkermansia* but not closely related taxa (**Fig. 1C, Supplementary Table S1)**. The genus has been linked to improved ICI response in several studies (*5–7*, *10*). The BRB group showed depletion of a few genera, including *Ruminiclostridium 5* (log2fc = −4.80, adjusted p-value = 0.049) and *Clostridium sensu stricto* 1 (log2fc = −8.21, adjusted p-value = 0.049), supporting specific effects of the intervention (**Fig. 1D**). This suggests that BRB supplementation may be a specific dietary intervention for enriching PIRMs.

**Fig. 1.**
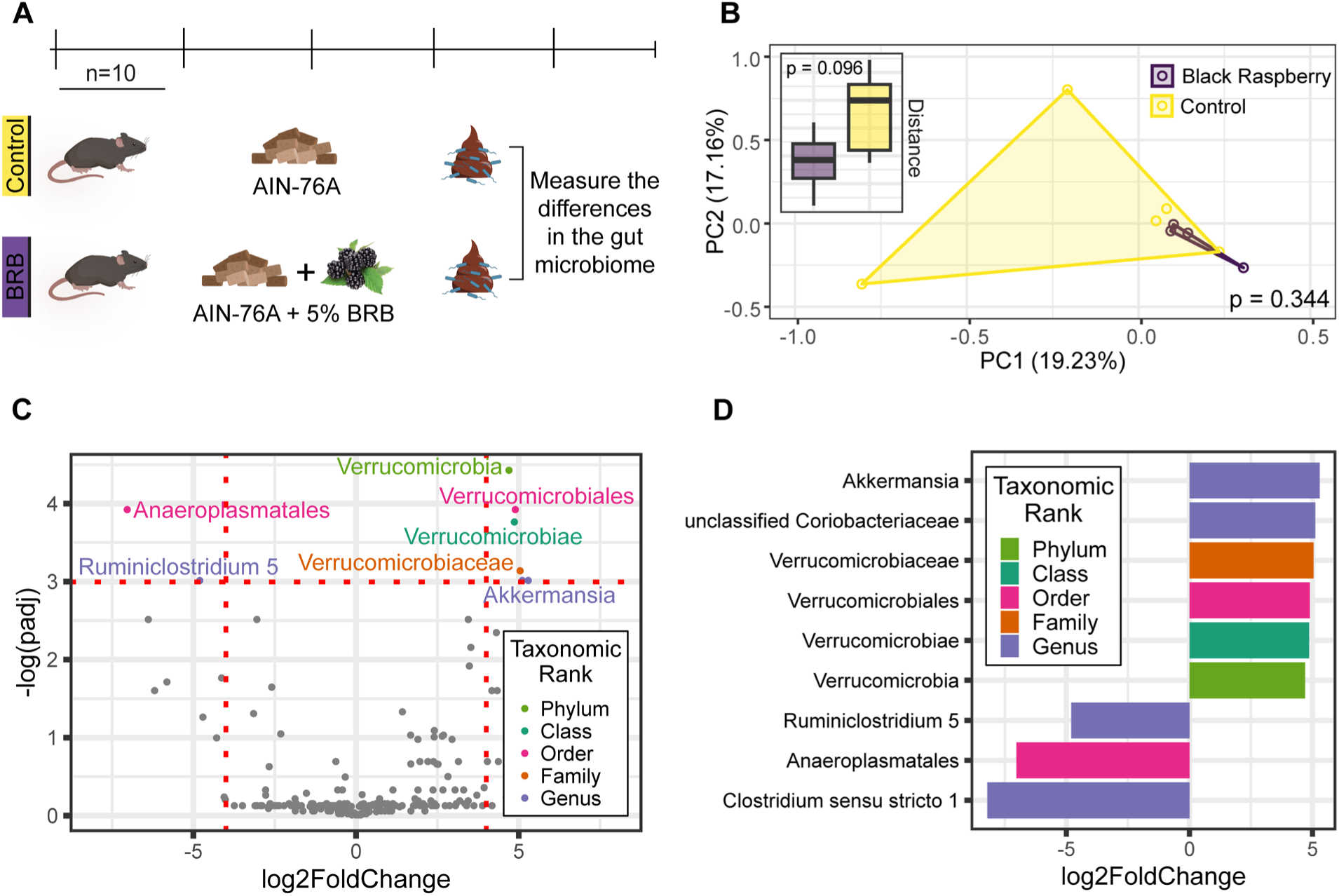
Black raspberry diet supplementation in a rat model enriches for gut PIRMs. (A) Schema of dietary supplementation studies in rat models fed with AIN-76A synthetic diet +/− 5% lyophilized black raspberry powder and the collection of gut microbiome samples. (B) Amplicon sequencing showed an increased abundance of *A. muciniphila* in the treatment group. (C) The change in microbiome composition was narrowly restricted to *A. muciniphila*, with the only other enriched taxonomic groups being from the same lineage.

### Human dietary intervention clinical trial in a cohort at high risk of developing lung cancer

Following the animal BRB diet intervention study results, we began a human clinical trial to explore whether similar changes in PIRMs could be obtained. The intervention delivered 340 mL of BRB nectar daily for four weeks (**Figure 2A**). The study population was recruited from a lung cancer screening clinic; they were cancer-free, but phenotypically similar to patients with lung cancer to maintain as much relevance to cancer as possible while reducing treatment-derived variation. We used a crossover design, where participants were randomized first to receive the intervention or a taste- and texture-matched placebo control. Participants drank their respective treatments twice daily for four weeks, followed by a 2-week washout. After washout, participants switched to the other treatment for another four weeks, making the total study duration ten weeks. Stool samples were collected before and after each intervention **(Fig. 2A)**. Overall, the composition of the participants’ gut microbiomes remained similar, with *Bacteroidetes* and *Firmicutes* predominating (**Fig. 2B**). We used mixed-effects models to identify the individual taxa enriched after only the BRB time point. We observed an increased abundance of several taxa, including *Blautia obeum* and *Agathobacter rectalis*. These taxa, as well as several other of the most significantly enriched organisms, are members of the family *Lachnospiraceae* (e.g., *Roseburia sp.* CAG 309*, Lachnospiraceae unclassified*, *Lachnospira pectinoschiza*). (**Fig. 2C, Supplementary Table S2**). The effect size of BRB was largest for *Roseburia sp.* CAG 309 (**Fig. 2D**).

**Fig. 2.**
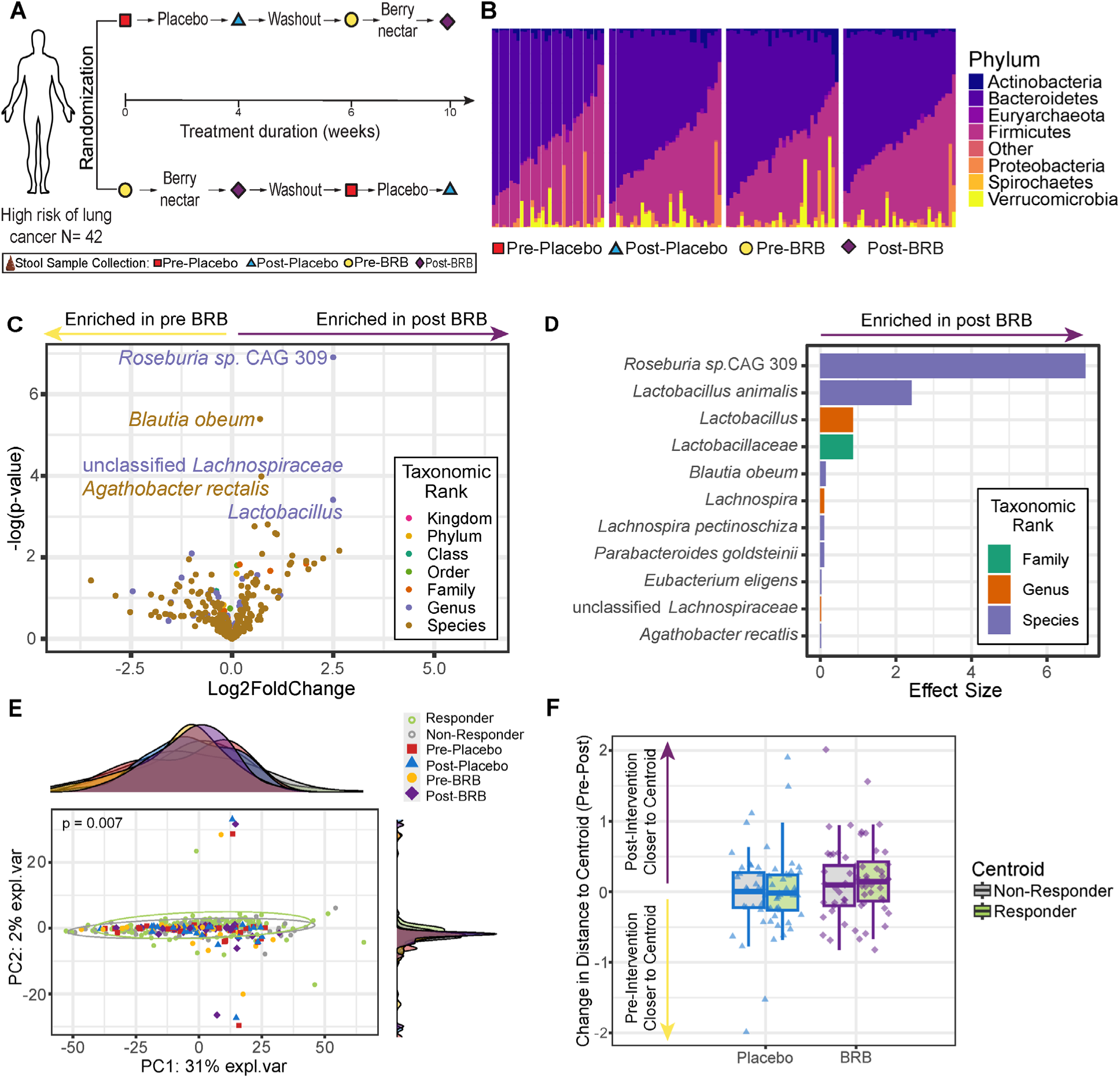
BRB nectar increases *Lachnospiraceae* abundance in the BEWELL human trial. **(A)** A crossover trial tested BRB nectar or placebo to participants for four weeks, with stool samples collected before and after each phase. **(B)** BRB treatment increased *Firmicutes* and decreased *Bacteroidetes* compared to placebo. **(C)** BRB significantly enriched *Lachnospiraceae* members, including *Blautia obeum* and *Agathobacter rectalis*. **(D)** *Roseburia sp CAG 309* showed the largest BRB-induced effect size. **(E)** Comparing the BEWELL gut microbiome communities to publicly available clinical trial data from cancer patients who responded or did not respond to ICIs showed no clear similarity to either group. However, after BRB supplementation, the total community was slightly more similar to a “Responder” type microbiome than pre-BRB or either placebo category **(F)**.

We next sought to put these changes into the context of cancer treatment outcomes. Previous studies have demonstrated the ability to predict whether an individual would respond to ICIs based on pre-treatment gut microbiome samples. We assembled publicly available data from five ICI clinical trials (n = 211) that assessed patients’ pre-treatment gut microbiome communities and related them to treatment response (*5*, *6*, *11–13*). As reported previously, the uniformly processed metagenomic sequencing data showed distinct clusters for responders and non-responders (adonis p-value = 0.007, **Fig. 2F**) (*5–7*). Overlaying the BEWELL trial data showed no major shifts in the gut microbiome communities, consistent with the relatively small number of altered microbes (**Fig 2B & C**). However, comparing the distance to the centroids of the non-responder or responder clusters showed that the post-BRB samples were more responder-like (**Fig. 2E**).

### Extending the human dietary intervention to cancer using preclinical models

Next, we sought to test whether the human trial’s dietary intervention modified the microbiome in a way that would affect the response to ICIs. We turned to a model system whereby a group of tumor-bearing mice were given a study participant’s microbiome from before the dietary intervention, and another group was given that person’s post-dietary intervention microbiome. We then monitored the effects of the different microbiomes on ICI response (**Fig. 3A**). We selected stool samples from study participants who exemplified the strongest changes across the cohort (i.e., those who showed large changes in the *Lachnospiraceae*, which had the most statistically significant enrichment by BRB), as well as two individuals who did not show these changes (**Fig. 3B**). As an additional control, we included stool from patients with lung cancer whose tumors responded well to ICIs or did not. Linear mixed effects models showed a significant negative effect on tumor volume of the gavage in the lung cancer patient whose tumor responded well to ICI treatment but had no significant association with the NR patient (**Fig. 3C**). Study participants with higher *Lachnospiraceae* in their post-BRB microbiome showed significantly smaller tumor volumes. Participants who did not show *Lachnospiraceae* showed no such effect. This suggests that the BRB-modified microbiomes, most strongly enriched for *Lachnospiraceae*, promoted tumor response to ICIs (**Fig. 3C**).

**Fig. 3.**
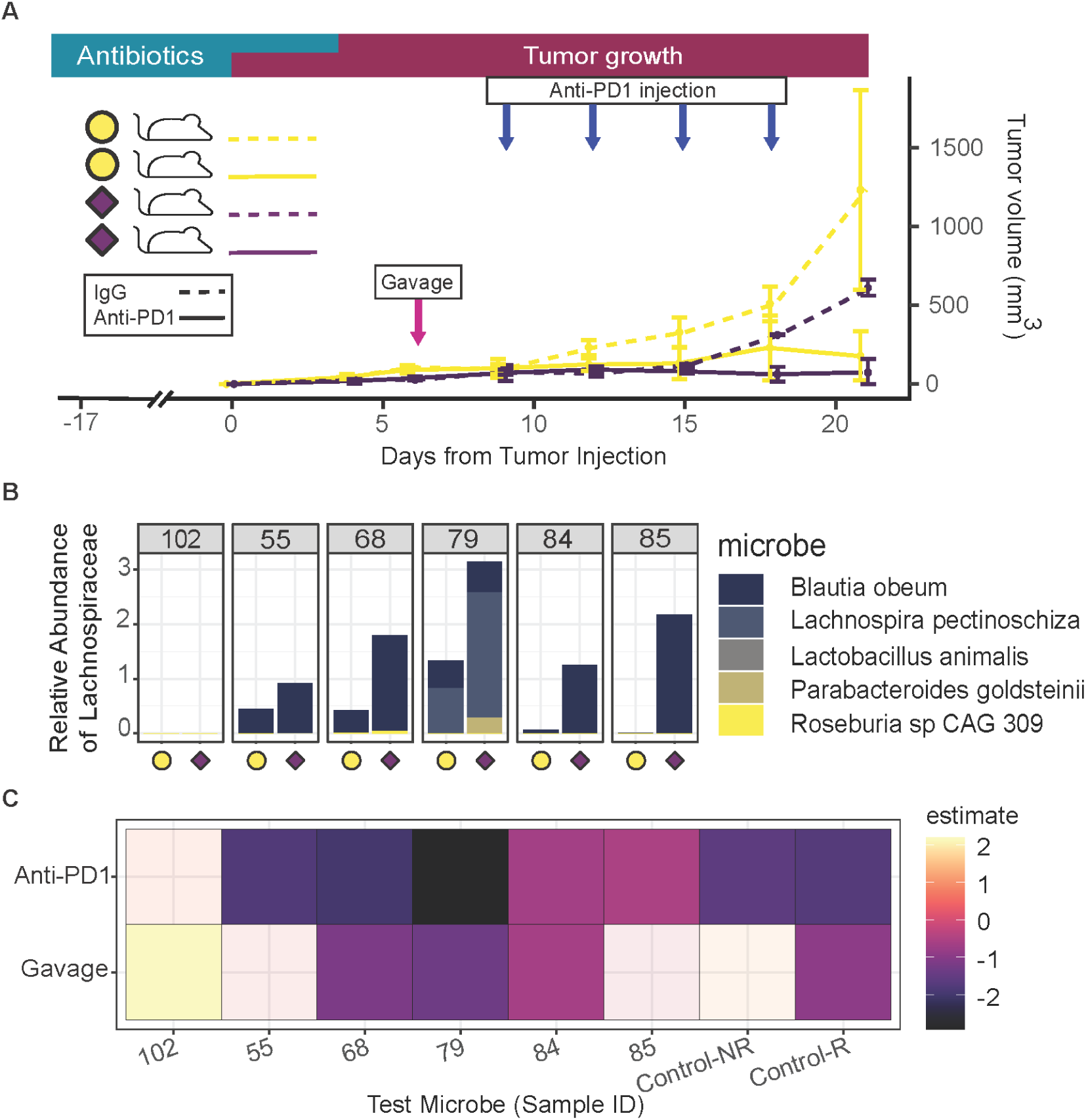
BRB-Modified Microbiomes Enhance ICI Response. **(A)** Tumor growth curves show significant tumor suppression in mice gavaged with post-dietary intervention microbiome and treated with anti-PD1 therapy compared to controls. **(B)** The abundance of *Lachnospiraceae* in samples gavaged into mouse models. **(C)** Study participants with higher *Lachnospiraceae* in their post-BRB microbiome showed significantly smaller tumor volumes. Participants who did not show *Lachnospiraceae* showed no such effect. Control-NR and Control-R microbiomes were from patients whose lung cancer was treated with ICIs and whose tumors did or did not respond to treatment.

### *Blautia* enrichment improves ICI response

Following the improvement in ICI response in mouse models that received high *Lachnospiraceae* samples (**Fig. 3B** & **C**), we aimed to evaluate whether *Blautia* alone could improve ICI response. We established a mouse model where one group received a fecal slurry from a representative pre-dietary intervention sample (Participant 85, **Fig. 4A**), while another group was gavaged with the same slurry supplemented with *Blautia*. All mice were injected with a cancer cell line and treated with anti-PD1. Monitoring tumor growth over time indicated that mice receiving the *Blautia*-supplemented slurry in conjunction with anti-PD1 treatment exhibited a marked reduction in tumor growth compared to those receiving the slurry alone (p-value= 0.012, **Fig. 4B**). Moreover, a higher proportion of mice in the *Blautia*-supplemented group remained tumor-free, emphasizing its potential to bolster therapeutic responses (**Fig. 4C**).

**Fig. 4.**
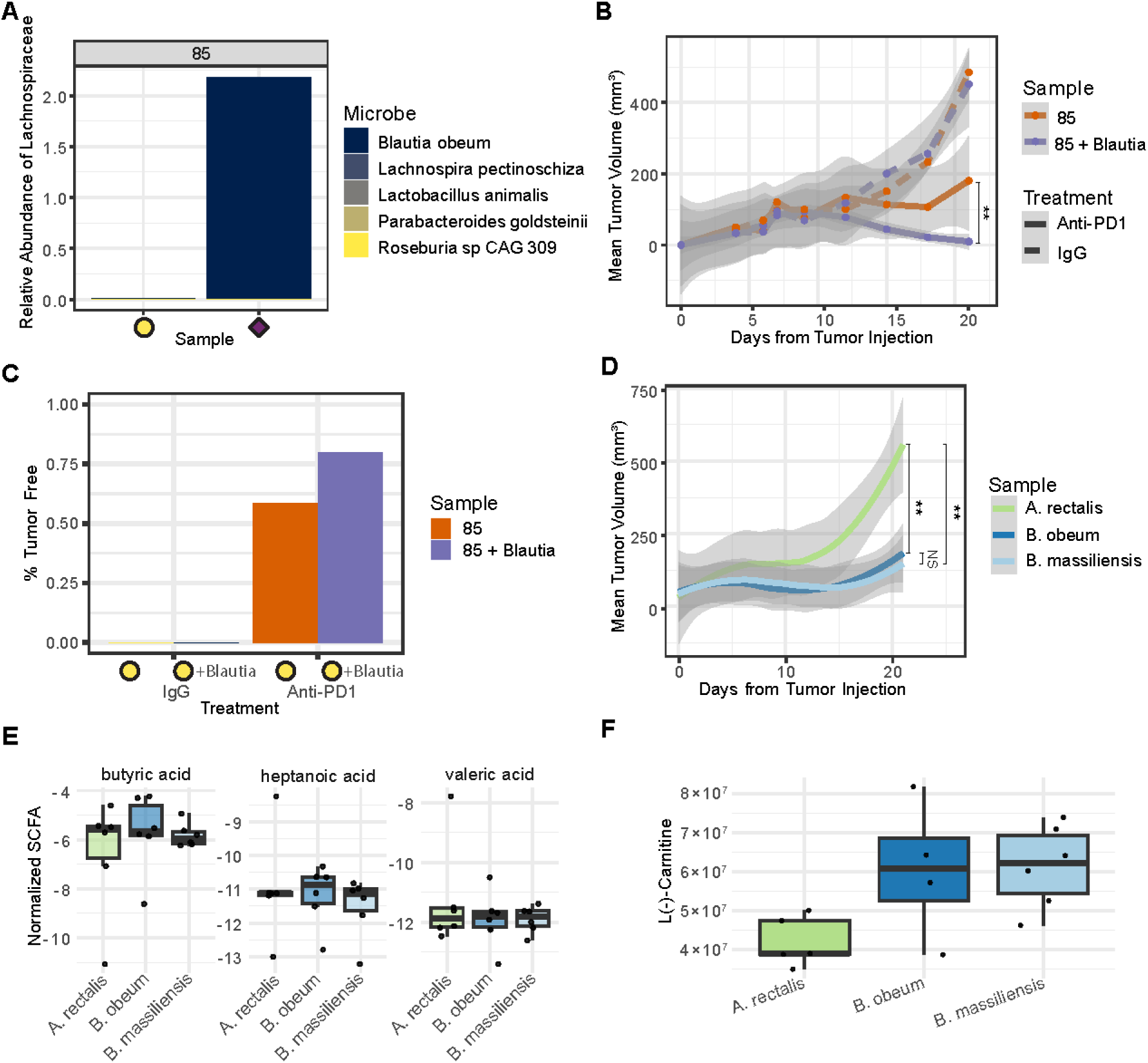
*Blautia* enrichment improves ICI response and alters metabolic profiles. (A) Mice gavaged with participant 85’s pre-BRB fecal slurry did not contain *Blautia*. (B) *Blautia*-supplemented mice showed reduced tumor growth. (C) More mice in the *Blautia* group were tumor-free. (D) *B. obeum* and *B. massiliensis* improved ICI response more than *A. rectalis*. (E) SCFA levels were unchanged across groups. (F) Elevated L-carnitine levels correlated with smaller tumors

To understand if the ICI effects we observed with *Blautia* are general across the *Lachnospiraceae*, we tested two closely related *Blautia* species (*obeum* and *massiliensis*) as well as a more distantly related family member that was enriched by BRB, *Agathobacter rectalis*. Results demonstrated that both *B. obeum* and *B. massiliensis* were more effective in slowing tumor growth than *A. rectalis*, suggesting that these microbes share the mechanism augmenting the benefits of immunotherapy (**Fig. 4D**).

Then we conducted a metabolomics analysis to assess the impact of microbial treatments on metabolic profiles. We measured short-chain fatty acids (SCFAs) like butyric acid, heptanoic acid, and valeric acid across different groups of mice treated with *A. rectalis*, *B. obeum*, and *B. massiliensis*. We observed no difference in the SCFA production across the groups (**Fig. 4E**), nor in other molecules that have been associated with ICI response, e.g., inosine or tryptophan derivatives (**Supplementary Fig. S1**). However, the mouse groups with smaller tumor volumes showed significantly greater L (−) carnitine levels (**Fig. 4F**).

## Discussion

In our study, BRB supplementation led to microbiome changes that may enhance ICI treatment outcomes. In rat feeding studies, BRB supplementation enriched for previously reported PIRMs like *Akkermansia*. In the human trial, a small BRB dietary intervention was sufficient to alter participants’ microbiomes and increase the abundance of the family *Lachnospiraceae*. When transferred to mouse models, these altered microbiomes led to improved tumor response to ICI treatment. Supplementation with *Blautia* alone enhanced ICI efficacy. Metabolomics suggests a role for increased L(−) carnitine in driving improved tumor responses to ICI treatment.

In recent years, the microbiome has been shown to play a causal role in the response to ICIs, leading many to study how best to alter the microbiome to improve treatment outcomes. Groups have explored the use of Fecal Microbiota Transplant (FMT), which has shown promise in several tumor types (*14–16*). Others have explored the use of single probiotics or microbial consortia (*17*), and several clinical trials are underway. We and others have pursued dietary interventions; here, we focus on polyphenols, but others have studied the effect of increased fiber (*9*) or specific fiber supplements like inulin (*18*) or pectin (*19*). These dietary approaches aim to provide a patient-accessible option for enhancing ICI efficacy through gut microbiome modulation.

High polyphenol dietary interventions have been used to modify health outcomes in several contexts. The source of the polyphenols are typically plants and include fruits (BRB (Rubus occidentalis), Camu camu (Myrciaria dubia) (*20*)), leaves (green tea (Camellia sinensis) (*21*, *22*), and seeds (walnuts (Juglans regia) (*23*)). A recent study by Messaoudene et al. (2022) demonstrated the potential of Camu camu to enhance antitumor activity and overcome resistance to ICI treatment through its effects on the gut microbiome (*20*). Previous work involving BRB has demonstrated broad anticancer properties. Chen et al. showed reduced tumorigenesis in a colon cancer mouse model fed BRB and suggested a mechanism involving demethylation of the gene Secreted frizzled-related protein 2 (SRFB2) (*24*). In clinical trials, BRB supplementation has led to reduced inflammatory cytokine circulation in patients with precancerous lesions or cancers of the oral cavity (*25*), esophagus (*26–30*), and colon (*31*, *32*). Here, we show a relatively small dietary intervention, without additional control of the diet, was sufficient to alter the gut microbiome. These findings underscore the potential of dietary interventions to improve cancer treatment outcomes, particularly in combination with ICI therapies.

The mechanism by which polyphenols alter health is expected to be highly varied but may be due to changes in microbial composition. We used preclinical models to relate the altered microbiome to treatment outcomes and showed that mice gavaged with the stool from individuals with BRB-enriched *Lachnospiraceae* had smaller tumor volumes. However, the altered microbes varied depending on the berry and conditions. When we fed BRB powder to rats, non-*Lachnospriaceae* PIRMs, including *Akkermansia,* were enriched. Messaoudene et al. showed that Camu camu berry increased the abundance of *Ruminococcus*. In our human trial, the BRB most strongly increased the abundance of *Lachnospiraceae* members.

*Lachnospiraceae* have been strongly associated with ICI response in multiple studies. McCulloch et al. (2022) analyzed microbial signatures across five melanoma cohorts, finding *Lachnospiraceae* most strongly linked with favorable outcomes to ICI treatment (*12*). In addition, Davar et al. (2021) also showed that FMT from donors whose tumors had responded to ICI treatment with high gut microbiome *Lachnospiraceae* abundance led to improved responses in PD-1–refractory melanoma patient recipients. The expected mechanism included increasing CD8+ T cell activation and modulating immunosuppressive myeloid cells (*15*). Studies in colorectal cancer provide further support for the benefits of *Lachnospiraceae*, with tissue-resident species like *Ruminococcus gnavus* and *Blautia producta* shown to promote immune surveillance by activating CD8+ T cells through the degradation of lyso-glycerophospholipids. This process restores CD8+ T cell function by counteracting the immunosuppressive effects of these metabolites (*33*). Lastly, a high abundance of *Lachnospiraceae* in colorectal cancer patients has been associated with elevated RNAseq-derived “immunoscores”, suggesting greater lymphocyte infiltration and a more favorable prognosis (*34*). Collectively, these studies provide strong correlative evidence for *Lachnospiraceae*’s role in enhancing ICI efficacy and its potential as a microbial biomarker for treatment response.

The precise mechanism by which *Lachnospiraeae* alter ICI response remains unclear. Several groups have reported the production of SCFA’s, notably butyrate, by *Lachnospiraceae* members, including *Roseburia* and *Eubacterium*. Higher levels of butyrate have been associated with improved ICI response in mouse and human studies, likely due to butyrate’s ability to enhance immune function by promoting the activation of cytotoxic T cells and NK cells. This modulation helps to improve immune surveillance and response to tumors, as butyrate has been shown to directly activate immune pathways such as NF-κB signaling in T cells (*8*, *35*, *36*). However, no increase in butyrate or other SCFAs was observed in the *Blautia*-supplemented mouse feces or blood. Other mechanisms include inosine and indole-3-aldehyde, but no differences in indole molecules correlated with tumor response in the mouse models. However, we did observe altered carnitine levels in mice gavaged with *Blautia*, leading to smaller tumor volumes, but not in mice gavaged with *Agathobacter rectale*, which showed larger tumor volumes. Carnitine appears to play a multifaceted role in animal biology, with its effects varying based on the context and cancer type. As a vital molecule for fatty acid transport into mitochondria, carnitine supports energy production, which is necessary for maintaining gut epithelial barrier function and intestinal homeostasis (*37*). However, certain cancers that rely on lipid metabolism may exploit carnitine to fuel their growth and progression. For instance, the carnitine palmitoyltransferase (CPT) system, crucial for fatty acid oxidation, has been identified as a potential target for anti-cancer therapies (*38*).

Our study highlights promising associations between BRB supplementation, microbiome enrichment, and improved ICI responses; however, it has some limitations. While BRB supplementation selectively enriched beneficial microbes like *Akkermansia* and *Lachnospiraceae*, the specific mechanisms driving improved ICI outcomes remain unclear. Although metabolomics revealed changes in L(−) carnitine levels, the role of carnitine in ICI efficacy is context-dependent, with both beneficial and potentially tumor-promoting effects reported.

## Conclusion

A BRB whole-food intervention shows promise as a means to modify the microbiome to improve ICI responses by selectively enriching beneficial microbes like *Lachnospiraceae* and modulating metabolic pathways. These findings contribute to the growing body of evidence supporting the role of the gut microbiome in shaping ICI outcomes. While our data suggest a role for metabolites such as L(−) carnitine in influencing ICI efficacy, further research is needed to unravel the underlying mechanisms and resolve conflicting reports regarding its impact. By offering a patient-accessible strategy, dietary interventions like BRB have the potential to complement existing ICIs and provide personalized treatment options. Moving forward, integrating microbiome modulation into ICI-based cancer care will require mechanistic studies, larger trials, and exploration of its applicability across diverse patient populations and cancer types. These efforts could pave the way for transformative advances in cancer treatment through microbiome-driven approaches.

## Data Availability

All data produced are available online at https://github.com/spakowiczlab/dmm21

## Acknowledgments

We would like to thank Angela Dahlberg, editor in The Ohio State University Division of Medical Oncology, for editing and proofreading this manuscript. We also thank the study participants for their effort and dedication.

## Funding

Pelotonia, clinical trial award (DS)

OSUCCC Division of Medical Oncology, seed grant (DS)

Association for the Study of Lung Cancer, Lung Cancer Foundation of America and the Bristol Meyers Squibb Foundation, Young Investigator in Immuno-Oncology Award (DS)

National Institutes of Aging grant 1K01AG070310 (DS)

The Ohio State University Wexner Medical Center and The Ohio State University College of Medicine Clinical Research Center/Center for Clinical Research Management

American Lung Association Innovator Award (DS)

## Author contributions

Conceptualization:

Methodology:

Investigation:

Visualization:

Funding acquisition:

Project administration:

Supervision:

Writing – original draft:

Writing – review & editing:

## Data and materials availability

## Supplementary Materials

Materials and Methods

Figs. S1 to S5

Tables S1 to S3

