## Supplemental materials for "Berry Supplement-Modified Human Microbiome Improves Immunotherapy Response in Mouse Models"

Science
AAAS

### Supplementary Materials for

#### Berry Supplement-Modified Human Microbiome Improves Immunotherapy Response in Mouse Models

Shiva Jahanbahkshi†, Amna Bibi†, Rebecca Hoyd, Caroline Dravillas, Nyelia Williams, Shiqi Zhang, Aaditya Pallerla, Shankar Suman, Joseph Amann, Mounika Goruganthu, Tamio Okimoto, Yangyang Liu, Marisa A. Bittoni, Ni Shi, Alvin Anand, Bailey Conrad, Lane Nevers, Kristen Heitman, Maxine Webb, Elizabeth M Grainger, Madison Grogan, Christian Quiles, Tong Chen, Carolyn Presley, Lang Li, Patrick Bradley, Yael Vodovotz, David P. Carbone, Steve K. Clinton, Jiangjiang Zhu, Daniel Spakowicz

**The PDF file includes:**

Materials and Methods
Figs. S1 to S3
Tables S1 to S4
References

#### Materials and Methods

Rat model feeding studies and 16S analysis

Gut microbiome samples were obtained from 10 male F344 rats (Harlan Sprague-Dawley (Indianapolis, IN, USA), equally split between control (AIN-76A synthetic diet) and treatment (AIN-76A synthetic diet containing 5% lyophilized black raspberry powder (BRB, Stokes Berry Farm)) groups. AIN-76 contains 20% casein, 0.3% d,l-methionine, 52% cornstarch, 13% dextrose, 5% cellulose, 5% corn oil, 3.5% AIN salt mixture, 1% AIN vitamin mixture, and 0.2% choline bitartrate (Dyets, Inc., Bethlehem, PA, USA). BRB was mixed into the diet (modified by reducing the concentration of cornstarch by 5% to maintain an isocaloric diet) for 25 min. with a Hobart mixer (Troy, OH, USA). At 35 weeks, stool pellets were collected, and all animals were euthanized by $CO_2$ asphyxiation and subjected to gross necropsy (Institutional Animal Care and Use Committee (IACUC) of The Ohio State University (OSU); Protocol # 2009A0054-R2). DNA was purified using a PowerFecal Pro kit (QIAGEN) with lysis by bead beating for 2 minutes on a PowerLyzer 24 (QIAGEN). Amplicon sequencing of the 16S v3-v4 rDNA from the samples was performed via Illumina MiSeq (2×250 bp). Reads were processed through the DADA2 pipeline (v1.16) (*1*) to generate amplicon sequence variant abundance tables and were classified compared to the SILVA database (v123) (*2*). Microbe abundances were compared between the two groups using a Kruskall-Wallis test.

Dietary Intervention Clinical Trial

We conducted a human intervention trial, the BEWELL Study (Black Raspberry nEctar Working to Prevent Lung cancer, NCT04267874), reviewed by the Ohio State University Comprehensive Cancer Center Institutional Review Board (2019C0089). We recruited 96 participants who were classified as being at high risk of developing lung cancer (eligibility criteria: >30 pack-year smoking history and 55-77 years old). This placebo-controlled, randomized, crossover trial examined the impact of 2 170-mL black raspberry (BRB) nectar drink boxes daily for 4 weeks. The drink contained 8% lyophilized BRB powder by weight, mixed with pectin, sugar, corn syrup, water, and citric acid as described previously (Gu et al. J Agric Food Chem 2015). The Pre- and post-dietary intervention gut microbiome, blood, and urine samples were collected. Participants collected gut microbiome samples at home using a Fisher Commode (Catalog No. 02-544-208, Thermo Fisher Scientific Inc, Waltham, MA USA), from which 500 mg of stool was collected into an OMR200 device (DNAgenotek, Toronto, Canada). Participants were instructed to keep the commode chilled and bring it to the clinic within 24 hours of the study visit.

Genomic DNA was purified by Powerfecal Pro (QIAGEN), and whole genome shotgun libraries were created by QIAseq FX (QIAGEN). Sequencing data were generated on a Novaseq 6000 platform. Microbiome abundance tables were obtained by processing the resulting FASTQs using MetaPhlAn3 using the option to include viruses in the database used for alignment.

Gut microbiome and ICI-response meta-analysis

We downloaded metagenomic data from the European Nucleotide Archive (ENA) for five publicly available datasets (*3–7*) (Accession numbers available in Supplemental table X). Metadata were collected from ENA and the corresponding NCBI BioProjects and available supplemental data. We filtered out non-metagenomic samples, repeat measurements, post-treatment samples, and samples lacking response information. The remaining samples were uniformly processed with BEWELL. Response categories were derived from the original study definitions based on RECIST or PFS. Microbe counts were normalized across studies using

MMUPHin (v3.20) (*8*) and clusters compared using adonis2 in the vegan package. Cluster centroid positions were calculated using the betadispr function, also in the vegan package, and the Euclidean distance was calculated to each point in the BE WELL dataset using dist in R. Code to regenerate all figures is available at https://github.com/spakowiczlab/dmm21.

Preclinical modeling with clinical trial gut microbiome

Six-week-old C57BL/6J mice were purchased from Jackson Laboratory and housed in BSL1 containment, and the acclimation period lasted for 1 week. Next, we depleted the gut microbiome with a 3-week antibiotic water regimen (*9*). The antibiotic solution was prepared at 1 mg/mL. Antibiotics with varying absorption levels were used. The high-absorption antibiotic cocktail included ampicillin, cefoperazone sodium salt, and clindamycin hydrochloride. The low-absorption antibiotic cocktail included ertapenem sodium, neomycin sulfate, and vancomycin hydrochloride. Mice remained on the antibiotic water regimen for 7 days, followed by 2 days of normal drinking water. Antibiotic water was also added to a small container of food pellets. Mice were visually inspected and weighed weekly for weight gain/loss and dehydration. Frozen gut microbiome samples from the Molecular Carcinogenesis and Chemo-prevention biobank (2015C0069, dietary intervention clinical trial) or the Total Cancer Care biobank (2013H0199, cancer patients treated with immunotherapy) were accessed via a protocol approved by the Ohio State University Comprehensive Cancer Center Institutional Review Board (2020C0131). Samples were thawed, and 1g was resuspended in 10 mL of sterile degassed phosphate-buffered saline (PBS). Degassed PBS was left in an anaerobic chamber for more than 12 hours. Mice were gavaged with 200 ul of stool slurry. Mouse colon cancer cells (mc38) were cultured, and $1e^6$ cells were injected subcutaneously in the flank of the mice. Nine days after cancer cell injection, mice were treated with BioXcell InvivoMAb anti-PD1 (5mg/kg; clone RMP1-14) or InVivoMab rat IgG2a isotype control (clone 2A3). Tumors were measured every 3 days by caliper, and stool pellets were collected. The mice were then euthanized 21 days after cancer cell injection. A cardiac puncture was conducted and placed on ice to be preserved for blood processing of p16. Tumors were extracted and placed in formalin or liquid nitrogen for immunohistochemistry (IHC) or 16S analysis. Tumors were, at times, kept fresh for flow cytometry processing. Euthanized mouse organs were surgically removed, including the spleen, duodenum, jejunum, and ileum, and placed in a fixative solution. The liver and colon were also collected and flash-frozen in liquid nitrogen. This animal study protocol was reviewed and approved by the OSU IACUC (Protocol # 2020A00000063-R1). Freshly extracted tumors were placed in Petri dishes and chopped using autoclaved blades. Chopped tumors were processed using a mouse tumor dissociation kit (Miltenyi Biotec).

16S data generation and processing

Amplicon sequencing of the 16S v3-v4 rDNA from the samples was performed via Illumina MiSeq (2×250 bp). Reads were processed through the DADA2 pipeline with the R package “dada2” (v1.12.1) to generate amplicon sequence variant abundance tables and were classified by comparison to the SILVA database (v123).

Fecal straight- and branched-chain fatty acids quantification

Fecal samples from mice (~20 mg each) were placed in screw-cap tubes with glass beads and homogenized using a Bead Homogenizer (BioSpec, Bartlesville, OK) in 20 µL of 10 mM 13C-labeled sodium butyrate (Cambridge Isotope Laboratories, Tewksbury, MA) and 180 µL of

50% aqueous acetonitrile. The homogenized samples were centrifuged at 14,000×g for 10 minutes at room temperature, and the resulting supernatant was mixed with 3-nitrophenylhydrazine (3NPH) and N-(3-dimethylaminopropyl)-N′-ethylcarbodiimide (v:v:v = 2:1:1). The mixture was incubated at 40°C for 30 minutes to produce 3NPH-fatty acid (FA) derivatives. A series of diluted standards was prepared for quantification as we reported before (*10*). The samples were analyzed using a Vanquish Liquid Chromatography system coupled with Q Exactive Mass Spectrometry (LC/MS) (Thermo Fisher Scientific, Waltham, MA). Separation of compounds was achieved on an Acquity UPLC CSH C18 column (1.7 µm, 2.1 × 100 mm, Waters, Milford, MA). Derivatized FAs were detected at their specific mass-to-charge ratios using electrospray ionization in negative mode. The mass spectra were acquired with a scan range of 60–900 m/z, a spray voltage of 2.75 kV, and an AGC target of 3e6. Quality control (QC) samples were run every ten biological samples to monitor instrument stability. Quantification results were calculated based on a standard curve and normalized to fecal weight.

Fecal and plasma LC/MS-based targeted metabolomics

Mouse fecal samples (~20 mg) were placed in screw-cap tubes containing glass beads and homogenized using a Bead Homogenizer (BioSpec, Bartlesville, OK) in 50 µL of $^{13}C^{15}N$-labeled amino acid mix (Cambridge Isotope Laboratories) and 250 µL of methanol. Mouse plasma samples (20 µL) were mixed with 50 µL of $^{13}C^{15}N$-labeled amino acid mix and 250 µL of methanol. Both fecal and plasma samples were stored at -20°C for 20 minutes to precipitate proteins, followed by centrifugation at 14,000 rpm for 15 minutes to collect the supernatant. A 200 µL aliquot of the supernatant was subjected to metabolomics analysis using an LC/MS system (Thermo Fisher Scientific). Separation of metabolites was performed on an Xbridge BEH Amide column (2.5 µm, 2.1 × 150 mm, Waters, Milford, MA, USA). Metabolites were detected in both positive and negative ion modes, with mass spectra acquired over a scan range of 60–900 m/z, a spray voltage of 2.75 kV, and an AGC target of 3e6. Pooled QC samples were injected every ten biological samples to ensure instrument stability. Peak alignment, integration, and compound annotation were conducted using Compound Discoverer 3.3 (Thermo Fisher Scientific, Waltham, MA, USA) with reference to an in-house database (*11*). Data files containing compound annotations and peak area information were filtered by a QC coefficient of variation < 0.25, and redundant metabolites were removed.

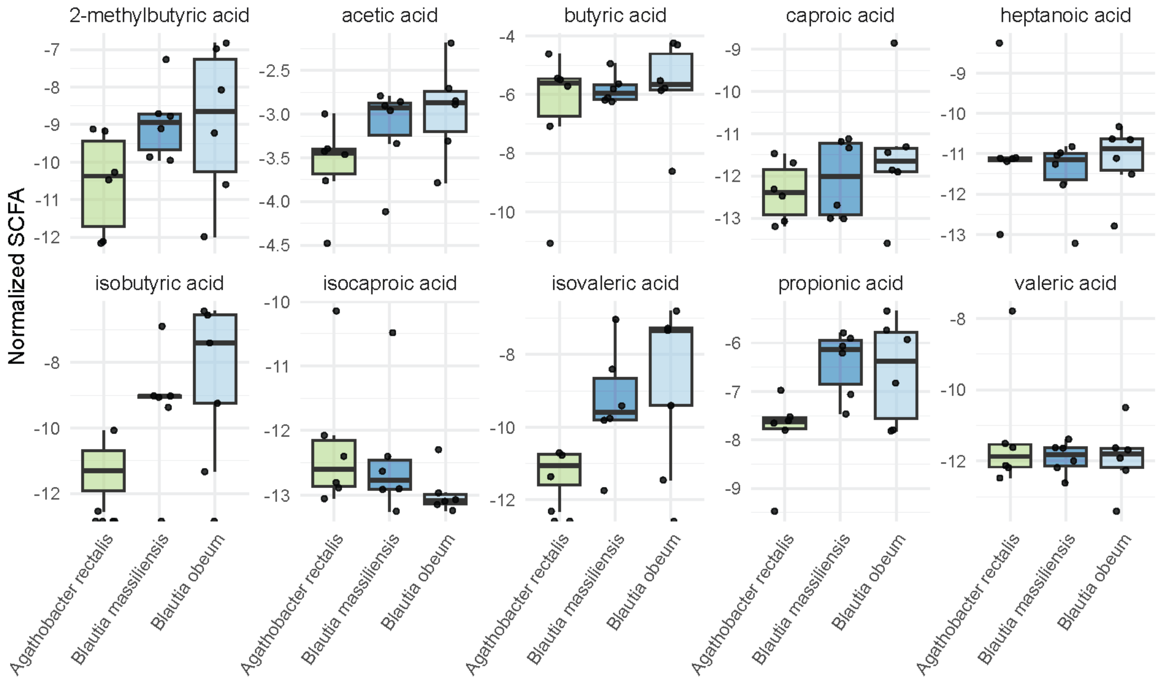


**Fig. S1. Metabolomic analysis of short-chain fatty acids in mice feces gavaged with *A. rectalis*, *B. massiliensis*, or *B. obeum*.** Mass spectrometer peak intensities, normalized to the weight of the stool pellet before extraction, for ten microbe-derived SCFAs are shown. There were no significantly different metabolites between the *Blautia* vs. *Agathobacter* groups.

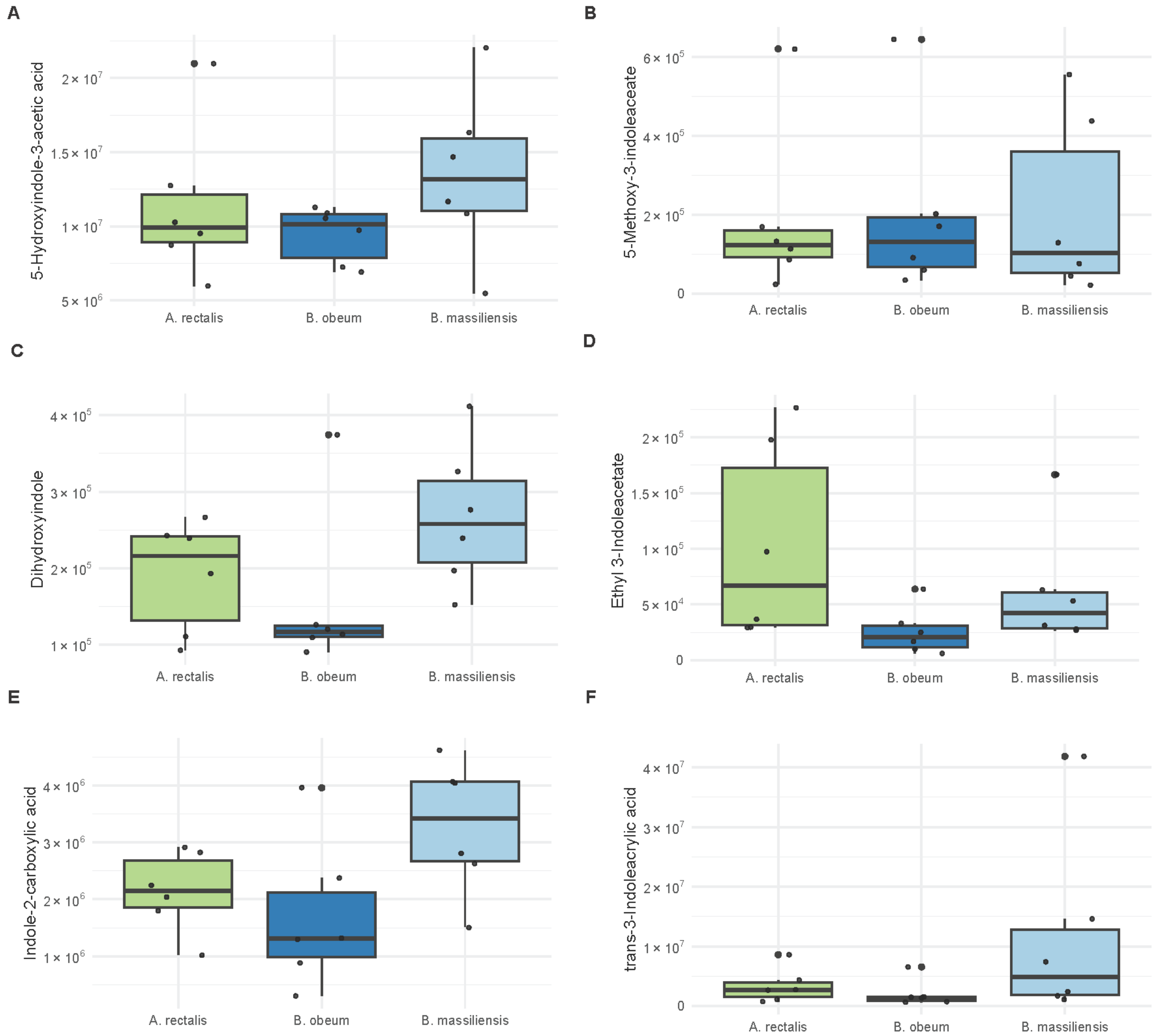


**Fig. S2. Metabolomic analysis of indole-containing molecules from mouse feces.**

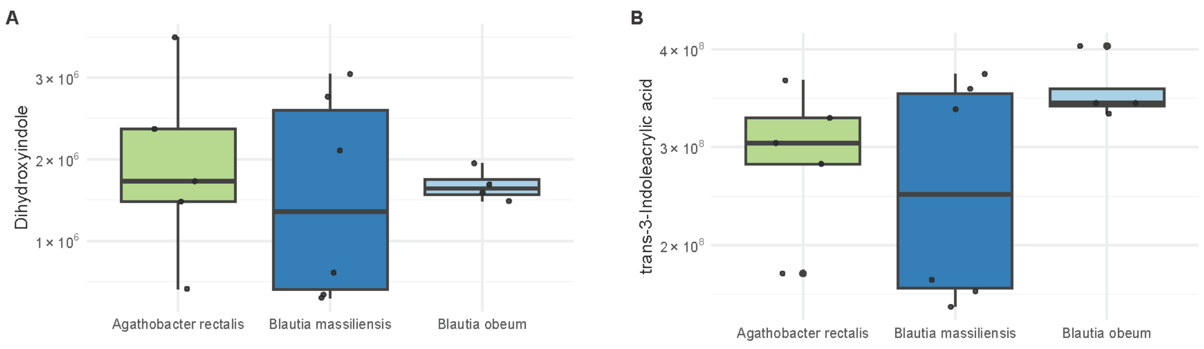


**Fig. S3. Metabolomic analysis of indole-containing molecules from mouse blood.**

**Table S1. Differential abundance analysis 16S data from rat feces following BRB supplementation.**

**Table S2. Differential abundance analysis metagenomics data longitudinal human stool samples throughout the BEWELL clinical trial.**

**Table S3. Accession numbers for the {respondersig} ICI-response meta-analysis.**

| Study | Study Accession | N | Responders | Non-Responders |
|---|---|---|---|---|
| **Frankel, 2017** | PRJNA397906 | 39 | 24 | 15 |
| **Gopalakrishnan, 2018** | PRJEB22893 | 25 | 14 | 11 |
| **Matson, 2018** | PRJNA399742 | 39 | 15 | 24 |
| **McCulloch, 2022** | PRJNA762360 | 94 | 68 | 26 |
| **Peters, 2020** | PRJNA541981 | 14 | 7 | 7 |

**Table S4. Longitudinal mixed-effects model results for the change in tumor volume predicted by ICI treatment and stool gavage.**
